# Association of Marital/Partner Status with Hospital Readmission Among Young Adults With Acute Myocardial Infarction

**DOI:** 10.1101/2023.06.20.23291664

**Authors:** Cenjing Zhu, Rachel P Dreyer, Fan Li, Erica S Spatz, César Caraballo, Shiwani Mahajan, Valeria Raparelli, Erica C Leifheit, Yuan Lu, Harlan M Krumholz, John A Spertus, Gail D’Onofrio, Louise Pilote, Judith H Lichtman

## Abstract

**Introduction:** Despite evidence supporting the benefits of marriage on cardiovascular health, the impact of marital/partner status on the long-term readmission of young acute myocardial infarction (AMI) survivors is less clear. We aimed to examine the association between marital/partner status and 1-year all-cause readmission, and explore sex differences, among young AMI survivors.

**Methods:** Data were from the VIRGO study (Variation in Recovery: Role of Gender on Outcomes of Young AMI Patients), which enrolled young adults aged 18-55 years with AMI (2008-2012). The primary end point was all-cause readmission within 1 year of hospital discharge, obtained from medical record, patient interviews, and adjudicated by a physician panel. We performed Cox proportional hazards models with sequential adjustment for demographic, socioeconomic, clinical and psychosocial factors. Sex-marital/partner status interaction was also tested.

**Results:** Of the 2,979 adults with AMI (2002 women [67.2%]; mean age 48 [interquartile range, 44-52] years), unpartnered individuals were more likely to experience all-cause readmissions compared with married/partnered individuals within the first year after hospital discharge (34.6% versus 27.2%, hazard ratio [HR]=1.31; 95% confidence interval [CI], 1.15-1.49). The association attenuated but remained significant after adjustment for demographic and socioeconomic factors (adjusted HR, 1.16; 95%CI, 1.01-1.34), and was not significant after further adjusting for clinical factors and psychosocial factors (adjusted HR, 1.10; 95%CI, 0.94-1.28). Sex-marital/partner status interaction was not significant (p=0.69). Sensitivity analysis using data with multiple imputation, and restricting outcomes to cardiac readmission yielded comparable results.

**Conclusions:** In a cohort of young adults aged 18-55 years, unpartnered status was associated with 1.3-fold increased risk of all-cause readmission within 1 year of AMI discharge. Further adjustment for demographic, socioeconomic, clinical and psychosocial factors attenuated the association, suggesting that these factors may explain disparities in readmission between married/partnered versus unpartnered young adults. Whereas young women experienced more readmission compared to similar-aged men, the association between marital/partner status and 1-year readmission did not vary by sex.

## Introduction

Despite an overall reduction in cardiovascular disease (CVD) prevalence and acute myocardial infarction (AMI) mortality,(1) rates of AMI hospitalization in young adults (≤55 years) have increased over the last two decades.(2) Hospital readmission remains frequent across all age groups of AMI survivors, with an overall 24% readmission rate within 90 days post-AMI.(3) In a recent study of US young adults with AMI, about one-third had at least 1 hospitalization in the year after discharge, and young women experienced more adverse events than men.(4) Risk profile for readmission among younger and older patients may be different, as suggested by a study using data from the 2013 National Radmission Database, where the effect of sex was more prominent in the younger age group.(3) Socio-demographic and psychosocial characteristics have been suggested to play important roles in predicting the risk of 1-year readmission for younger adults with AMI,(5) yet little is known about the impact of marital/partner status on their long-term risk of readmission.

Marriage has long been known to offer cardiovascular health benefits, including its association with lower risk of AMI incidence,(6) in-hospital and long-term mortality,(7–10) and recurrent events.(11–13) Committed relationships that are not based on formal legal unions, such as domestic partners and common-law marriages may also convey benefits, but are less commonly described in prior studies.(7,9,10,12) Moreover, prior research has largely focused on older populations, been conducted in foreign countries, and has not explored readmission beyond the first month of discharge.(11,12) There is a paucity of data on the impact of marital/partner status on the long-term readmission outcomes of younger AMI patients. In addition, although evidence suggest that women may not benefit from marriage to the same extent as men regarding mortality outcomes(14,15), less is known about whether there are sex differences in the degree of “protection” conferred by marriage/partnership in a younger population with AMI and as assessed by hospital readmission.

To address this gap in knowledge, we examined the association between marital/partner status and all-cause and cardiac readmission within 1 year of hospital discharge among a cohort of AMI survivors 18-55 years of age in the United States. A secondary aim was to explore potential subgroup differences in the association by sex.

## Materials and Methods

### Study Population

We used data from the VIRGO study (Variation in Recovery: Role of Gender on Outcomes of Young AMI Patients), the largest prospective, multicenter cohort study designed to understand factors associated with adverse outcomes in younger adults (≤55 years) with AMI.(16) Between August 21, 2008 and May 1, 2012, a total of 2,979 participants were recruited from 103 US hospitals using a 2:1 female-to-male enrollment design. The methodology of VIRGO has been described elsewhere.(16) In brief, eligible participants had elevated cardiac enzymes (troponin or creatine kinase, with at least one of these biomarkers >99^th^ percentile of the upper reference limit) at the recruiting center within 24 hours of admission, and presented with other evidence supporting the diagnosis of AMI, including either symptoms of ischemia or electrocardiogram changes indicative of new ischemia (new ST-T changes, new or presumably new left bundle branch block, or the development of pathological Q waves). Patients were excluded if their elevated cardiac markers were due to a complication of elective coronary revascularization, or their AMI was caused by physical trauma. Individuals were ineligible if they were incarcerated, did not speak English or Spanish, or were unable to provide informed consent or be contacted for follow-up.

This study followed the Strengthening the Reporting of Observational Studies in Epidemiology (STROBE) reporting guideline (**Appendix** in the **Supplement**). De-identified data were used for the current study. Institutional review board approval was obtained at each participating institution, and individuals provided written informed consent for their study participation.

### Assessment of Marital/Partner Status and Other Covariates

Baseline data were collected by medical chart abstraction and standardized in-person interviews administered by trained personnel during the index AMI admission. Marital/Partner status was collected during patient enrollment interview through a question of “Which best describes your current marital status” and was categorized into “married/partnered” (having a response of “married” or “living as married/living with partner” or “unpartnered” (having a response of “divorced”, “separated”, “widowed”, or “single”). In a secondary analysis, “unpartnered” status was further classified into “divorced/separated”, “widowed”, or “single”.

Demographic factors included sex (male/female), age (year, continuous), and self-reported race (non-Hispanic white/non-Hispanic black/Hispanic/other [ie, American Indian or Alaska Native and Asian or Pacific Islander]). Socioeconomic factors included education level, financial strain, employment status, and health insurance. Education level was categorized into less than high school, some high school, and more than high school.

Financial strain was defined as having “just enough to make ends meet” or “not enough to make ends meet” (versus having some money left over) based on individuals’ response to the question “In general, how do your finances usually work out at the end of the month”.

Clinical characteristics considered in our study included cardiac risk factors (hypertension, dyslipidemia, diabetes, obesity, current smoker, and alcohol abuse), medical history (prior CVD [AMI, percutaneous coronary intervention, coronary artery bypass grafting, angina, heart failure, stroke, transient ischemic attach or peripheral artery disease], renal dysfunction, and history of chronic obstructive pulmonary disease [COPD]), and disease severity (type of AMI [ST-elevation myocardial infarction/non-ST-elevated myocardial infarction], ejection fraction <40%, and hospital length of stay).

Psychosocial factors, including depression, low social support, and high stress burden, were assessed at baseline by validated measures or questionnaires respectively. Depressive symptoms were measured using the 9-item version of the Patient Health Questionnaire,(17) with an overall score of 10 or more indicating depression. Social support was measured using the 5-item Enhancing Recovery in Coronary Heart Disease Social Support Inventory.(18) Low social support was defined as a score of 3 or less on at least 2 Social Support Instrument items and a total score of 18 or less.(19) High stress burden was captured by answering “Fairly often” or “Very often” to the 14-item Perceived Stress Scale(20) question “In the last month, how often have you felt nervous and stressed?”

### Collection and Adjudication of Hospital Readmission

The primary end point of this study is all-cause readmission within 1 year of hospital discharge. During the 1-year follow-up period, hospital readmissions were identified by the research coordinators at each site from medical record and self-report. The VIRGO adjudication process was supported through the use of a custom-developed Research Electronic Data Capture external module.(21) Adjudications of all-cause and cardiac readmission were completed by five physicians and an advanced practice registered nurse at Yale University who received extensive training and clear guidelines. Detailed process has been described elsewhere.(5) In sensitivity analysis, we restricted outcomes to cardiac readmission and results remained consistent.

### Statistical Analysis

Baseline characteristics were compared between married/partnered versus unpartnered participants, overall and by sex, using χ^2^ test for dichotomous variables and Wilcoxon’s rank-sum test for continuous variables that don’t follow a normal distribution. All variables had minimal missing values (<5%) except for financial strain (11%). In sensitivity analysis, multiple imputation by chained equations was applied to generate 10 imputed datasets on which estimates were calculated and pooled by the Rubin’s rule.(22) Modeling was performed both with and without the imputation of missing values and because results were almost identical, we reported the complete case analysis in the main paper and presented the results from multiple imputation in **eTable 1** in the **Supplement**.

Time to readmission was compared between married/partnered and unpartnered groups using the log-rank test. To examine the independent association of marital/partner status with all-cause readmission over the subsequent 1 year after AMI, multivariable Cox proportional hazard models sequentially adjusted for 4 domains of covariates including demographics (age, sex, race), socioeconomic factors (education level, financial strain, employment status, and health insurance), clinical characteristics (cardiovascular risk factors, medical history and disease severity), and psychosocial factors (depression, low social support, high stress burden). Covariates adjusted in the multivariable models were selected using a combination of clinical judgement and insights from previous literature,(5,23,24) with a detailed variable selection procedure described elsewhere.(5) Two-way interaction between marital/partner status and sex was also tested in the fully-adjusted model using the Wald χ^2^ test. The proportional hazard assumption was checked by Schoenfeld residual with a global test p>0.05 indicating no violation.

Due to competing risk from non-cardiac readmissions, in the sensitivity analysis, Fine-Gray competing risk model were applied to examine the association between marital/partner status with the cumulative incidence of cardiac readmission within 1-year post discharge. The same set of covariates considered in the Cox model were adjusted for in the Fine-Gray model.

All statistical analyses were conducted using R (Version 1.4.1106), with 2-tailed tests for statistical significance indicated by P=0.05. Data analysis was performed from February to July 2022.

## Results

Among the 2,979 participants 18-55 years of age (median age 48 years, interquartile range 44-52), 42.8% were unpartnered (781 divorced/separated, 91 widowed, 423 single; 47% for women and 37.2% for men). Baseline characteristics are presented in **Table 1**.

**Table 1.** Baseline characteristics of the study participants by marital/partner status and sex.

|  | Married/Partnered (N=1675) |  |  | Unpartnered (N=1299) |  |  | P-value |
| --- | --- | --- | --- | --- | --- | --- | --- |
|  | Men<br>(n=610) | Women<br>(n=1065) | p-value | Men<br>(n=362) | Women<br>(n=937) | p-value | (unpartnered<br>vs.<br>married/partn<br>ered) |
| *Demographics* |  |  |  |  |  |  |  |
| Age (Median [q1, q3]) | 48 [44, 52] | 48 [44, 52] | 0.898 | 48 [43, 51] | 48 [44, 52] | 0.085 | 0.259 |
| Race |  |  | 0.319 |  |  | <0.001 | <0.001 |
| Non-Hispanic white | 476 (78.0%) | 813 (76.3%) |  | 252 (69.6%) | 532 (56.8%) |  |  |
| Non-Hispanic black | 53 (8.7%) | 134 (12.6%) |  | 52 (14.4%) | 275 (29.3%) |  |  |
| Hispanic | 43 (7.0%) | 64 (6.0%) |  | 39 (10.8%) | 87 (9.3%) |  |  |
| Other | 38 (6.2%) | 54 (5.1%) |  | 19 (5.2%) | 43 (4.6%) |  |  |
| *Socioeconomic factors* |  |  |  |  |  |  |  |
| Education level |  |  | 0.148 |  |  | 0.005 | 0.007 |
| Less than high school | 10 (1.6%) | 18 (1.7%) |  | 6 (1.7%) | 24 (2.6%) |  |  |
| Some high school | 220 (36.1%) | 435 (40.8%) |  | 183 (50.6%) | 383 (40.9%) |  |  |
| More than high school | 376 (61.6%) | 605 (56.8%) |  | 169 (46.7%) | 523 (55.8%) |  |  |
| Financial strain |  |  | <0.001 |  |  | 0.029 | <0.001 |
| Yes | 340 (55.7%) | 686 (64.4%) |  | 254 (70.2%) | 729 (77.8%) |  |  |
| No | 212 (34.8%) | 279 (26.2%) |  | 55 (15.2%) | 97 (10.4%) |  |  |
| Missing | 58 (9.5%) | 100 (9.4%) |  | 53 (14.6%) | 111 (11.8%) |  |  |
| **Employment status** |  |  | <0.001 |  |  | 0.075 | <0.001 |
| Unemployed | 124 (20.3%) | 434 (40.8%) |  | 145 (40.1%) | 440 (47.0%) |  |  |
| Employed | 483 (79.2%) | 631 (59.2%) |  | 217 (59.9%) | 495 (52.8%) |  |  |
| **Health insurance** |  |  | 0.964 |  |  | <0.001 | <0.001 |
| No | 104 (17.0%) | 187 (17.6%) |  | 137 (37.8%) | 246 (26.3%) |  |  |
| Yes | 504 (82.6%) | 874 (82.1%) |  | 221 (61.0%) | 690 (73.6%) |  |  |
| ***Clinical factors(cardiac risk factors and medical history, disease severity)*** |  |  |  |  |  |  |  |
| **Hypertension** | 378 (62.0%) | 659 (61.9%) | 0.999 | 249 (68.8%) | 687 (73.3%) | 0.264 | <0.001 |
| **High cholesterol** | 563 (92.3%) | 889 (83.5%) | <0.001 | 340 (93.9%) | 787 (84.0%) | <0.001 | 0.997 |
| **Diabetes** | 152 (24.9%) | 379 (35.6%) | <0.001 | 107 (29.6%) | 417 (44.5%) | <0.001 | <0.001 |
| **Obesity** | 300 (49.2%) | 545 (51.2%) | 0.735 | 164 (45.3%) | 562 (60.0%) | <0.001 | 0.013 |
| **Physical inactivity** | 163 (26.7%) | 361 (33.9%) | 0.009 | 132 (36.5%) | 373 (39.8%) | 0.575 | <0.001 |
| **Current smoker** | 210 (34.4%) | 338 (31.7%) | 0.529 | 80 (22.1%) | 263 (28.1%) | 0.091 | <0.001 |
| **Alcohol abuse** | 282 (46.2%) | 287 (26.9%) | <0.001 | 173 (47.8%) | 266 (28.4%) | <0.001 | 0.994 |
| **Prior cardiovascular disease** | 189 (31.0%) | 348 (32.7%) | 0.775 | 128 (35.4%) | 367 (39.2%) | 0.448 | 0.003 |
| **Renal dysfunction** | 41 (6.7%) | 111 (10.4%) | 0.040 | 42 (11.6%) | 142 (15.2%) | 0.254 | <0.001 |
| **COPD** | 31 (5.1%) | 130 (12.2%) | <0.001 | 31 (8.6%) | 153 (16.3%) | 0.002 | <0.001 |
| **Type pf AMI** |  |  | <0.001 |  |  | <0.001 | 0.417 |
| STEMI | 351 (57.5%) | 501 (47.0%) |  | 212 (58.6%) | 417 (44.5%) |  |  |
| NSTEMI | 259 (42.5%) | 564 (53.0%) |  | 150 (41.4%) | 520 (55.5%) |  |  |
| **EF<40%** |  |  | 0.259 |  |  | 0.033 | 0.333 |
| Yes | 60 (9.8%) | 132 (12.4%) |  | 48 (13.3%) | 79 (8.4%) |  |  |
| No | 535 (87.7%) | 898 (84.3%) |  | 304 (84.0%) | 830 (88.6%) |  |  |
| **Length of stay (Median [q1, q3])** | 3 [2, 4] | 3 [2, 4] | 0.178 | 3 [2, 4] | 3 [2, 5] | 0.02 | 0.009 |
| ***Psychosocial factors*** |  |  |  |  |  |  |  |
| **Depression** | 108 (17.7%) | 362 (34.0%) | <0.001 | 103 (28.5%) | 390 (41.6%) | <0.001 | <0.001 |
| **High stress burden** | 211 (34.6%) | 554 (52.0%) | <0.001 | 156 (43.1%) | 531 (56.7%) | <0.001 | <0.001 |
| **Low social support** | 82 (13.4%) | 172 (16.2%) | 0.341 | 130 (35.9%) | 253 (27.0%) | 0.007 | <0.001 |
| ***1-year readmission outcomes*** |  |  |  |  |  |  |  |
| **All-cause readmission** | 124 (20.3%) | 331 (31.1%) | <0.001 | 97 (26.8%) | 352 (37.6%) | 0.001 | <0.001 |
| **Cardiac readmission** | 94 (15.4%) | 228 (21.4%) | 0.001 | 78 (21.5%) | 241 (25.7%) | 0.293 | 0.002 |

During the first year of follow-up, 904 patients had ≥1 all-cause readmission (50.3% were married), 641 had ≥1 cardiac readmission (50.2% were married). Overall, compared to married/partnered individuals, those who were unpartnered had higher risk of all-cause readmission throughout the first year of recovery (**Figure 1A**). When further stratified by sex, married male had the lowest risk, while unmarried female had the highest (**Figure 1B**).

**Figure 1.**
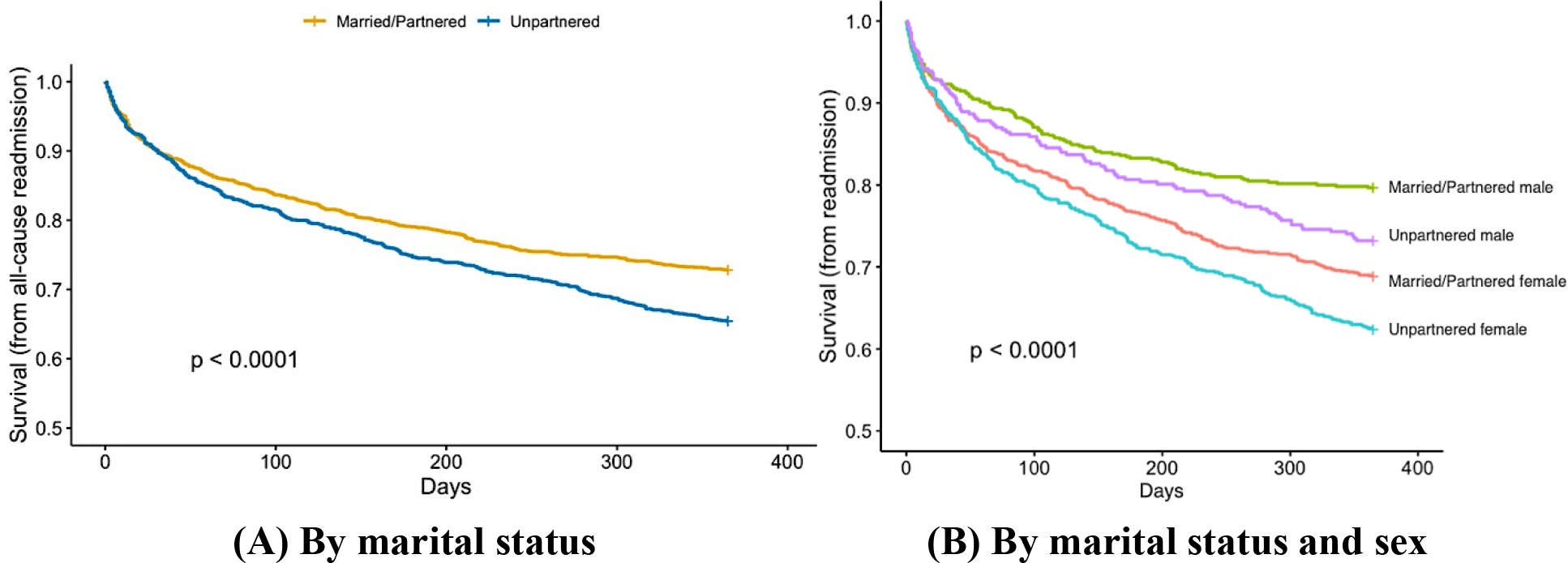
Kaplan Meier curves for time to all-cause readmission.

In multivariable analyses, compared with being married/partnered, being unpartnered was associated with a 24% higher risk of all-cause readmission after adjustment for demographic factors (age, sex and race) (aHR, 1.24; 95%CI, 1.08-1.42). The association attenuated after further adjusting for socioeconomic factors (education, financial strain, employment, insurance) (aHR, 1.16; 95%CI, 1.01-1.34), and further attenuated and became not statistically significant after adjusting for clinical factors (aHR, 1.11; 95%CI, 0.96-1.29) and psychosocial factors (aHR, 1.10; 95%CI, 0.94-1.28). Variables that were significant in the fully adjusted model include female sex, age, Hispanic race, financial strain, unemployment, diabetes, prior CVD, COPD, total length of stay, and depression. Details of the model output can be found in **Table 2**.

**Table 2.** Multivariable Cox regression models output.

|  | **Model1**<br>**(R<sup>2</sup>=4.7%)** | **Model2**<br>**(R<sup>2</sup>=9.1%)** | **Model3**<br>**(R<sup>2</sup>=15%)** | **Model 4**<br>**(R<sup>2</sup>=16%)** |
| --- | --- | --- | --- | --- |
| **Marital status (Unpartnered vs. Married/Partnered)** | 1.24 (1.08-1.42) * | 1.16 (1.01-1.34) * | 1.11 (0.96-1.29) | 1.10 (0.94-1.28) |
| ***Demographics*** |  |  |  |  |
| Female sex | 1.52 (1.31-1.78) * | 1.43 (1.21-1.69) * | 1.36 (1.14-1.62) * | 1.26 (1.05-1.51) * |
| Age, year | 0.99 (0.98-1.00) | 0.99 (0.98-1) | 0.98 (0.97-0.99) * | 0.98 (0.97-0.99) * |
| Race (ref: non-Hispanic white) | - | - | - | - |
| Non-Hispanic black | 1.22 (1.04-1.44) * | 1.18 (0.98-1.4) | 1.15 (0.95-1.38) | 1.19 (0.98-1.44) |
| Hispanic | 0.66 (0.50-0.88) * | 0.70 (0.52-0.95) * | 0.64 (0.47-0.88) * | 0.62 (0.45-0.87) * |
| Other race/ethnicity | 0.84 (0.61-1.17) | 0.89 (0.63-1.26) | 0.87 (0.61-1.25) | 0.91 (0.63-1.32) |
| ***Socioeconomic factors*** |  |  |  |  |
| Education (ref: less than high school) |  | - | - | - |
| Some high school |  | 0.91 (0.55-1.51) | 1.04 (0.61-1.78) | 0.97 (0.56-1.68) |
| More than high school |  | 0.86 (0.52-1.44) | 1.00 (0.58-1.70) | 0.93 (0.53-1.61) |
| Financial strain |  | 1.41 (1.15-1.72) * | 1.32 (1.08-1.63) * | 1.33 (1.07-1.64) * |
| Unemployment |  | 1.49 (1.30-1.72) * | 1.30 (1.11-1.54) * | 1.30 (1.10-1.52) * |
| No health insurance |  | 1.23 (1.04-1.47) * | 1.12 (0.93-1.34) | 1.14 (0.95-1.37) |
| ***Clinical factors (Cardiac risk factors, medical history, and disease severity)*** |  |  |  |  |
| Hypertension |  |  | 1.05 (0.88-1.25) | 1.01 (0.84-1.21) |
| High cholesterol |  |  | 1.13 (0.89-1.42) | 1.08 (0.85-1.37) |
| Diabetes |  |  | 1.31 (1.12-1.54) * | 1.33 (1.13-1.56) * |
| Obesity |  |  | 0.89 (0.77-1.04) | 0.88 (0.75-1.02) |
| Physical inactivity |  |  | 1.08 (0.93-1.25) | 1.05 (0.9-1.23) |
| Current smoking |  |  | 1.07 (0.91-1.27) | 1.11 (0.93-1.31) |
| Alcohol abuse |  |  | 0.96 (0.82-1.13) | 0.98 (0.83-1.16) |
| Prior cardiovascular disease |  |  | 1.23 (1.06-1.44) * | 1.22 (1.03-1.43) * |
| Renal dysfunction |  |  | 1.19 (0.96-1.47) | 1.23 (0.99-1.52) |
| COPD |  |  | 1.35 (1.11-1.65) * | 1.32 (1.07-1.62) * |
| STEMI |  |  | 1.00 (0.87-1.16) | 1.04 (0.89-1.21) |
| Ejection Fraction <40% |  |  | 0.92 (0.73-1.16) | 0.89 (0.7-1.13) |
| Total length of stay |  |  | 1.03 (1.02-1.05) * | 1.03 (1.01-1.05) * |

| *Psychosocial factors* |  |  |  |  |
| --- | --- | --- | --- | --- |
| Depression |  |  |  | 1.35 (1.13-1.61) * |
| Low social support |  |  |  | 0.97 (0.81-1.16) |
| High stress burden |  |  |  | 1.07 (0.91-1.26) |

\*p<0.05 indicating statistical significance.
Note: Model 1 adjusted for demographics; Model 2 adjusted for demographics and socioeconomic factors; Model 3 adjusted for demographics, socioeconomic, and clinical factors. Model 4 adjusted for demographic, socioeconomic, clinical and psychosocial factors. Covariates were pre-selected based on prior literature and clinical implications. Interaction between marital/partner status and sex was tested and was not significant in the fully adjusted models (p=0.628). Fully adjusted model does not violate the proportional hazard assumption (global p>0.05).

In the fully adjusted model, the two-way interaction between marital/partner status and sex was not significant (p=0.628). To provide additional information on the direction of the interaction, the fully adjusted models were also stratified by sex. Details of the sex-specific models output can be found in **eTable 2** in the **Supplement**.

Sensitivity analysis using data with multiple imputation, and restricting outcomes to cardiac readmission yielded comparable results (**eTable 1** and **eTable 3** in the **Supplement**). In the fully adjusted model with imputed data, no health insurance was also associated with all-cause readmission. When restricting outcomes to cardiac readmission using the Fine-Gray model, financial strain and depression were no longer significant in the fully adjusted model.

In a secondary analysis further classifying unpartnered participants into divorces/separated, widowed, and single subgroups, similar pattern was found among divorced/separated individuals as compared to the overall unpartnered group; widowed individuals had the highest risk of all-cause readmission, yet the association was not statistically significant due to the small size of widowed participants in current study; no association was found among single individuals. (**Table 3**)

**Table 3.** Association between marital/partner status subgroup and all-cause readmission.

|  | Model 1 | Model 2 | Model 3 | Model 4 |
| --- | --- | --- | --- | --- |
| **Married/Partnered (1675)** | ref | ref | ref | Ref |
| **Unpartnered (1299)** | 1.24 (1.08-1.42) * | 1.16 (1.01-1.34) * | 1.11 (0.96-1.29) | 1.10 (0.94-1.28) |
| **Divorced/Separated (781)** | 1.26 (1.08-1.46) * | 1.20 (1.02-1.41) * | 1.17 (0.98-1.38) | 1.15 (0.96-1.37) |
| **Widowed (91)** | 1.39 (0.99-1.95) | 1.29 (0.91-1.84) | 1.09 (0.76-1.57) | 1.04 (0.72-1.51) |
| **Single (423)** | 1.17 (0.96-1.43) | 1.06 (0.85-1.31) | 1.00 (0.80-1.26) | 1.00 (0.80-1.28) |

\*p<0.05 indicating statistical significance.
Note: Model 1 adjusted for demographics; Model 2 adjusted for demographics and socioeconomic factors; Model 3 adjusted for demographics, socioeconomic, and clinical factors. Model 4 adjusted for demographic, socioeconomic, clinical and psychosocial factors. Covariates were pre-selected based on prior literature and clinical implications.

## Discussion

In a cohort of young adults with AMI in the United States, we found a 1.3-fold higher readmission rate in unpartnered compared to married/partnered individuals. The association between marital/partner status and 1-year all-cause readmission attenuated and remained significant after adjusting for demographic and socioeconomic factors, but was not significant after further adjusting for clinical and psychosocial factors. Sex difference was not evident in the fully adjusted model.

Prior research generally supported improved survival and fewer recurrent events in married individuals compared to their unmarried counterparts within 1-year post AMI.(7–13) However, only a few studies investigated the impact of marital/partner status on post-event health outcomes beyond mortality among AMI survivors, with younger patients being underrepresented.(11–13) Our study addresses this important knowledge gap using adjudicated readmission data from a large nationwide cohort of young adults with AMI in the United States. Compared to an Isreali study of AMI pateints with a mean age of 64 years,(11) which found no unadjusted association between marital status and 30-day readmission (17.8% among married, 19.1% among nonmarried, p=0.28), our study showed a higher unadjusted all-cause readmission rate within 1 year after AMI among unpartnered individuals compared to married/partnered. Differences in the results could be due to a longer follow up time in our study, and different participant characteristics (eg, younger age, different culture).

Previous studies that reported an independent association between marital status and AMI outcomes have not generally accounted for socioeconomic or psychosocial factors in their analyses.(11–13) However, mounting evidence has demonstrated that these factors are more powerful predictors of adverse outcomes following AMI than physical health indicators, especially in a younger population.(5) Our finding of the association being attenuated after adjusting for socioeconomic status and clinical factors supported the roles of both clinical (ie, diabetes, CVD history, COPD, length of stay) and socioeconomical components (ie, financial and employment status) in explaining the complicated relationship between marriage/partnership and health outcomes. Further, these findings align with prevailing theories that explain the mechanism of marriage protection. First, the marriage selection theory suggests that healthier people may be more likely to get and stay married.(25) In our study we also observed poorer health at baseline, including clinical, behavioral and psychosocial factors, among unpartnered compared to married/partnered individuals. Second, the social causation theory centered on the health benefits from spousal support with regard to treatment adherence, lifestyle changes, as well as greater socioeconomic resources, which make healthy behaviors affordable.(26,27) In our study, while further adjusting for psychosocial factors didn’t substantially change the results, findings provided a more comprehensive risk factor profile for young adults with AMI, where unpartnered individuals were more likely to have depression, low social support, and high stress burden at baseline. Our results lend support to marriage selection and social causation theories in a younger population with AMI, but should be interpreted with caution since our exploratory analysis can only generate associational evidence. Future research is encouraged to include a formal mediation analysis to understand the complex relationship and potential causal pathway associated with these findings.

Studies on sex differences in the impact of marital/partner status on cardiovascular outcomes have yielded mixed findings, with the majority of prior work supporting a greater marital benefit for men than women.(6,10,13,15) On the contrary, our study did not find such a sex difference in the association in a younger population. The potential mechanism for such findings is likely to be the offset of biological and psychosocial effects. Physiologically, women were protected against heart disease by sex hormones such as estrogen that reduces circulatory levels of harmful cholesterol, testosterone that increase the concentrations of low-density lipoprotein and inflammatory markers that affect atherosclerosis and stroke progression.(28) From a psychosocial perspective, however, women had distinct vulnerabilities, including unique sources of psychosocial stress and distrimination, increased perceived stress during adulthood, and twofold greater lifetime prevalence of depression and anxiety disorders compared with men.(29–31) This was also evident in the current study that unmarried women had the highest likelihood to have depression and high stress burden. Taken together, while our study found the impact of marital/partner status being equal for younger men and women, it might involve a mixture of biological and psychosocial effects that warrant further investigation.

### Study Implications

This study adds to the understanding of the association between marital/partner status and readmission outcomes up to 1 year post event in younger adults with AMI. Readmission presents a complex interaction between patients, community, environment, and healthcare system, and is an important measure indicating health outcomes and disease burden. Usually perceived as a demographic variable, marital/partner status adds an important dimension of social support and is also an easily attainable indicator. Clinicians may consider incorporate patients’ marital/partner status along with other socio-demographic factors into risk assessment and decision-making to create a more patient-centered practice. Our study may also inform potential interventions based on the social and psychological context of younger adults with AMI. For example, support groups or secondary prevention programs could widen participation of unpartnered individuals to improve their psychosocial well-being and recovery.

### Limitations

Limitations of this study merit discussion. Self-reported readmission were validated with retroactive chart review but misclassification bias may still be present. Although our study included an extensive array of demographic, socioeconomic and clinical factors, residual confounding due to unmeasured characteristics that differ by marital/partner status may still bias the results. Our modeling approach generated only associational evidence instead of causation, therefore findings should be interpreted with caution. Participants enrolled in the VIRGO study may not reflect those who did not enroll in the study or hospitalized at other institutions.

## Conclusions

Compared to young adults with AMI who are married/partnered, unpartnered individuals had 1.3 times higher risk of all-cause readmission within 1 year after hospital discharge. The association attenuated yet remained significant after adjusting for demographic and socioeconomic factors, but was not significant after further adjusting for clinical and psychosocial factors. Sex difference was not evident in the association. Further study is needed to explore causal relationships.

## Data Availability

All supporting data of this study are available upon reasonable request from Dr. Yuan Lu.

## Acknowledgements

None.

## Sources of Funding

The VIRGO study was supported by a 4-year National Heart, Lung, and Blood Institute grant (No. 5R01HL081153). Dr Dreyer is supported by an American Heart Association (AHA) Transformational Project Award (#19TPA34830013). This project was additionally supported by a Canadian Institutes of Health Research project grant (PJT-159508).

## Disclosures

Dr. Spertus discloses providing consultative services on patient-reported outcomes and evidence evaluation to Alnylam, AstraZeneca, Bayer, Merck, Janssen, Bristol Meyers Squibb, Edwards, Kineksia, 4DT Medical, Terumo, Cytokinetics, Imbria, and United Healthcare. He holds research grants from Bristol Meyers Squibb, Abbott Vascular and Janssen. He owns the copyright to the Seattle Angina Questionnaire, Kansas City Cardiomyopathy Questionnaire, and Peripheral Artery Questionnaire and serves on the Board of Directors for Blue Cross Blue Shield of Kansas City. No other authors report having any other disclosures to report.

## Supporting Information Captions

**Appendix STROBE Statement Checklist**

**eTable 1. Multivariable Cox regression models output using multiple imputed data eTable 2. Sex-specific Cox regression model output**

**eTable 3. Multivariable Fine-Grey models (cardiac readmission)**

## Notes

### Competing Interest Statement

I have read the journal's policy and the authors of this manuscript have the following competing interests: Dr. Spertus discloses providing consultative services on patient-reported outcomes and evidence evaluation to Alnylam, AstraZeneca, Bayer, Merck, Janssen, Bristol Meyers Squibb, Edwards, Kineksia, 4DT Medical, Terumo, Cytokinetics, Imbria, and United Healthcare. He holds research grants from Bristol Meyers Squibb, Abbott Vascular and Janssen. He owns the copyright to the Seattle Angina Questionnaire, Kansas City Cardiomyopathy Questionnaire, and Peripheral Artery Questionnaire and serves on the Board of Directors for Blue Cross Blue Shield of Kansas City. No other authors report having any other disclosures to report.

### Funding Statement

The author(s) received no specific funding for this work.

### Author Declarations

Institutional Review Board approval was obtained at each participating institution, and patients provided informed consent for their study participation including baseline and follow-up interviews.

